# Muscle morphology and architecture of the medial gastrocnemius between typically developing children with different ancestral backgrounds

**DOI:** 10.1101/2023.01.10.23284392

**Authors:** F. Walhain, R. Chin A Fat, M. Declerck, L. Bar-On, A. Van Campenhout, K. Desloovere

## Abstract

**Introduction:** Muscle ultrasonography is frequently used to improve the understanding of musculoskeletal impairments in children with cerebral palsy. So far, most studies on muscle morphology and architecture have included typically developing children and children with cerebral palsy with similar ancestry, being mainly Caucasian. Less is known about differences in muscle morphology and architecture between children with different ancestral backgrounds. Therefore, the aim of this study was to compare muscle morphology and architecture of the medial gastrocnemius and Achilles tendon of Surinamese typically developing children from African, South Asian and Southeast Asian descent.

**Method:** This explorative cohort study included 100 typically developing children identified as Maroon (Ghana, African descent), Hindustani (India, South Asian) or Javanese (Indonesia, Southeast Asian), aged 5 to 10 years. A conventional B-mode 2D ultrasound was used to define anatomical cross-sectional area (aCSA), fascicle length and pennation angle (architectural parameters). The muscle belly length, volume and physiological cross-sectional area (pCSA), as well as the tendon length (morphological parameters) were defined using 3D freehand ultrasound, which combines B-mode 2D ultrasound with 3D motion tracking. Muscle and tendon lengths were normalized to the total muscle tendon unit (MTU) lengths and fascicle lengths to muscle belly lengths, while volume, aCSA and PCSA were normalized to body mass. One-way Anova with post hoc t-tests were used to investigate differences between the ancestral groups. A two-way repeated measures Anova was used to define whether the extensibility of the muscle tendon, belly and fascicle differed between ancestral groups for the three conditions, i.e. when applying 0Nm, 1Nm and 4Nm ankle dorsiflexion torque.

**Results:** The ancestral subgroups included 34 Hindustani, 34 Javanese and 32 Maroon children. Normalized belly length was 11% shorter in Maroon and 7% shorter in Hindustani children compared to Javanese children (*p* = <0.001 and *p* = 0.002, respectively). Normalized fascicle length of Javanese children was 23% longer compared to Maroon and 11% longer compared to Hindustani children (*p* < 0.001 and *p* = 0.010, respectively). Normalized muscle volume was significantly higher in Javanese compared to Hindustani children (*p* = 0.002). The normalized aCSA was higher in Javanese children compared to the Maroon children (*p* = 0.008), while pCSA was higher for Maroon children compared to the Hindustani children (*p* = 0.007). The pennation angle of the fascicle with the deeper aponeuroses was larger in Maroon compared to Javanese children (*p* = 0.015). There were no differences in the extensibility of the muscle belly, tendon and fascicle between ancestral groups.

**Discussion:** Ancestry-specific reference data of the morphology of the medial gastrocnemius and Achilles tendon are needed when investigating altered muscle morphology in children with cerebral palsy. The current study showed differences in morphology (muscle belly-, tendon length and muscle volume, aCSA) and architecture (pCSA, fascicle length and deeper pennation angle) between children with different ancestry. These differences were most pronounced for Javanese compared to Maroon or Hindustani children. Future studies should report the ancestral background when describing muscle morphology and architecture of children and ancestral specifications in normative databases should be included.

## INTRODUCTION

Muscle ultrasonography is increasingly used to improve the understanding of musculoskeletal impairments in children with spastic Cerebral Palsy (SCP) (Williams *et al*., 2021). During growth, adaptations in skeletal muscles, i.e., an increase in muscle length and anatomical cross-sectional area (aCSA), resulting in increasing muscle volume, occur in both typically developing (TD) children and children with SCP (Benard *et al*., 2011; Modlesky and Zhang, 2019; Bell *et al*., 2021; Handsfield *et al*., 2022). However, these increases are lower in children with SCP compared to TD children (Barber *et al*., 2016), while muscle tendons were found to be longer (Barrett and Lichtwark, 2010). For a good understanding of the musculoskeletal impairments in children with SCP, proper normative reference data of muscle morphology in TD children are important.

Most of the current studies on muscle morphology and architecture in children with SCP compared to TD children have been published in North America, Europe, the UK, Australia, and Korea (Williams *et al*., 2021). While the ancestry of the enrolled children of these studies was not systematically mentioned, it may be assumed that data have been primarily reported for Caucasian children. Less is known about differences in muscle morphology between children with different ancestral backgrounds, such as African, East Asian and South Asian descent.

To the best of our knowledge, only two previous studies compared muscle morphology and architecture between children of different ancestry (Kunimasa *et al*., 2022). Song et al. (2002) reported smaller total muscle volume of both the upper and lower limbs in East Asian and Caucasian children compared to children of African descent (Song *et al*., 2002). This finding was not confirmed by Kunimasa *et al*. (2022), who recently reported smaller muscle thickness for the medial gastrocnemius (MG) in Kenyan (African) children compared to Japanese (East-Asian) children (Kunimasa *et al*., 2022). The latter study also observed shorter fascicle and longer Achilles tendon lengths, with no differences in pennation angle, in the Kenyan compared to Japanese children (Kunimasa *et al*., 2022). Most of these morphological and architectural differences were already observed at an age of 4 years. Due to the limited number of studies on ancestry-specific paediatric muscle morphology with contradicting results, there is a need for more data on muscle morphology parameters of TD children with different ancestral backgrounds.

Most previous studies that compared muscle morphology and architecture between ancestral groups, as well as between TD with SCP children of one specific ancestral group, focused only on one or a minimal selection of outcome parameters. However, an assessment of all relevant macroscopic parameters is required for a good understanding of the differences between groups. For example, morphological parameters, such as muscle volume or thickness, should be combined with architectural parameter, such as fascicle length, to derive the physiological cross-sectional area (pCSA). Furthermore, fascicle length should be accompanied by data on muscle belly and tendon length to understand the potential contributions to altered joint range of motion. Moreover, the MG lengths may be assessed at different ankle joint positions (i.e., when stretched by applying a specific joint torques), to improve the understanding of the compliance of the muscle and tendon in TD children. Integrated assessments that combine all morphological and architectural parameters may help to understand differences between ancestral groups and altered muscle properties in children with SCP.

The scarce results reported on ancestry-specific muscle morphology and architecture question the validity of pooling muscle morphology and architecture data of children with different ancestral backgrounds and suggest the need for an ancestral-specific reference database. This is especially relevant for regions with a population including different ancestral backgrounds, such as Suriname, where the research on children with SCP inherently includes children of mainly Indian (South Asian), Ghana (African) and Indonesian (Southeast Asian) descent. It is crucial to know the differences between children from different ancestral populations, before comparing muscle morphology and architecture of SCP and TD children.

Therefore, the aim of this explorative cohort study was to compare the muscle morphology and architecture of the MG of a large group of Surinamese TD children from African, South Asian and Southeast Asian descent, from 5 to 10 years old. The ancestral-specific morphological and architectural features for TD children were delineated and the comprehensive reference paediatric database is put available online to be used for future studies *(link to dataset will be added when the study is published)*.

## METHOD

### Study design and participants

This exploratory cohort study included TD children aged 5 to 10 years old. The ancestral background of the child was identified as Maroon (Ghana, African descent), Hindustani (India, South Asian) or Javanese (Indonesia, Southeast Asian) if at least three of the grandparents had the same ancestral background, while all other cases were categorized as ‘mixed’ group (Krishnadath *et al*., 2015). For homogeneous variation of age and sex in each age group, we aimed for an equal age and gender distribution per ancestral group. Children were included if at least three of the grandparents were categorized as Javanese, Hindustani or Maroon. Children were excluded in case of 1) mixed ancestral background, 2) having a history of neurological problems or developmental disorders, 3) previous lower leg musculoskeletal injuries, such as fractures or 4) doing organized sports three times or more in a week.

Ethical approval was given by the Medical Ethical Committee of the Ministry of Health (reference VG 003-17). Approval to perform the measurements at schools was given by the Ministry of Education, Science and Culture (NB/mz/Ag. 5653). Informed consent was obtained in writing from the parent/guardian of all children as well as the children’s verbal consent before inclusion.

### Recruitment and sample size determination

Recruitment and measurements of the children were performed from March 2021 to April 2022. Almost all children were recruited via primary schools and seven children were recruited via family and friends of co-workers of the Faculty of Medical Sciences of the Anton de Kom University of Suriname. All children were living in the capital city of Suriname, Paramaribo.

For sample size calculation, the effect size was estimated using the available literature data on differences in muscle mass, measured by dual energy X-ray absorptiometry, in children with different ancestral backgrounds from United States of America (effect size: 0.94 for girls and 0.41 for boys (Silva *et al*., 2010)) and the available literature data for the differences in adult Achilles tendon length among adults (effect size: 0.80 (Kunimasa *et al*., 2014) and 1.3 (Refsdal, 2017)). These previous studies suggest an averaged large effect size of 0.86. Based on a sample size calculation (independent t-test – power 0.80, alpha 0.0167 (corrected for 3 groups), effect size 0.86, G-power version 3.1.9.6) each ancestral group should include 30 children. We finally aimed to recruit 30 to 35 children per ancestral population, including 5 children for each of the 6 age-year categories, from 5 to 10 years old.

### Data collection and processing

In total, 5 schools gave permission for the measurements. At these schools, informed consent forms, as well as questionnaires on ancestral background of the grandparents, history of neurological or developmental disorders and frequency of weekly sport activities were handed out to the parents of the children that could meet the inclusion criteria. If children met de inclusion criteria and the parents and child agreed to participate, they were measured at school. For 7 children who were recruited via family and friends of co-workers of the Faculty of Medical Sciences, the measurements were performed at the Human Motion Laboratory at the Faculty of Medical Sciences of the Anton de Kom University of Suriname. The left and right leg were randomly selected by use of a coin toss. Anthropometric data were collected before the ultrasound assessment, while plantar flexor strength was assessed afterwards. Children lay prone during the collection of muscle morphological and architectural data and were instructed to relax while reading a book or watching a video. In case of body segment movements or muscle contraction of the participant, the acquisition was repeated. The duration of the measurement session lasted 45 minutes per child. All data were collected and processed by one assessor (first author, FW), who was trained and experienced in conducting clinical assessments, had 4 years of experience in performing ultrasound measurements, and was assisted by at least one physiotherapy student or colleague. During the data processing, anonymously name files for ancestry were used to ensure that FW was blinded for the ancestral background of the child.

### Anthropometric measurements

Anthropometric measurements included height, body weight, maximal passive ankle dorsiflexion, calf circumference and lower leg length *(for more details, supplementary I*). The position of the knee and ankle joint during all ultrasound conditions was measured using a universal goniometer (7” Baseline Plastic goniometer, Fabrication Enterprises). The ankle position was recorded with an interval of five degrees for each different condition, representing a plantarflexion position with a negative angle and a dorsiflexion with a positive angle.

### Muscle morphology and architecture outcomes

The morphological and architectural parameters were assessed in resting position (RP) of the ankle joint, i.e., with the foot hanging down freely at the end of a triangular pillow (*supplementary II, Figure 1a*). A conventional B-mode 2D ultrasound was used to define architectural parameters, namely the aCSA, fascicle length and (deeper and superficial) pennation angle *(for more details, supplementary II)*. To define the morphological parameters, namely muscle volume, pCSA, belly length, tendon length and MTU length, a previously described 3DfUS technique (Cenni *et al*., 2016) which combines B-mode 2D ultrasound with 3D motion tracking, was used. The ultrasound collection, the 3D reconstructions of the muscle and tendon, as well as all processing of the data, were performed with the open source STRADWIN software *(for more details, supplementary II)*.

**Figure 1:**
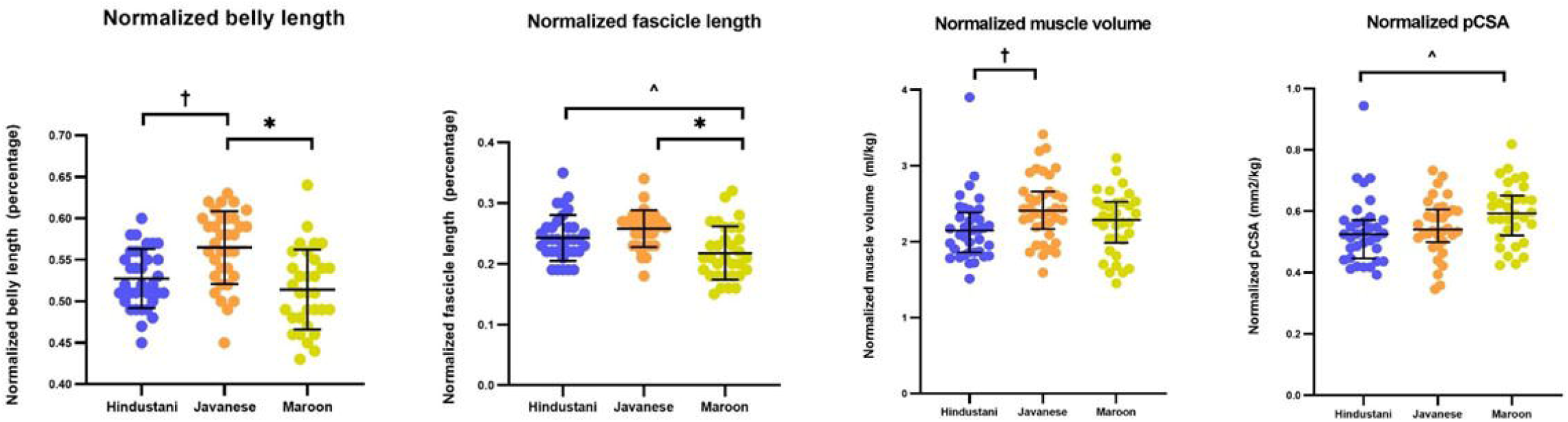
Individual data points for each participant (coloured), grouped by ancestry, for normalized belly length, fascicle length and muscle volume and physiological cross-sectional area in resting position. Mean and standard deviation (normalized belly and fascicle length) and median with interquartile range (normalized muscle volume and physiological cross-sectional area) are given (in black) for each ancestry. Group differences *(p* < 0.05) are displayed with; * for significant difference between Maroon and Javanese children; ^ for significant difference between Maroon and Hindustani children; † for significant difference between Hindustani and Javanese children.

Muscle volume and the aCSA and pCSA were normalized to body mass and muscle tendon and muscle belly length were normalized to muscle-tendon unit (MTU) length. Fascicle length was normalized to muscle belly length, since muscle belly length was different between ancestral groups.

### Extensibility

To assess the extensibility of the muscle tendon, belly and fascicle, the muscles and tendons were assessed in three other conditions. Therefore, the foot was placed in a custom-made set-up (*supplementary III, Figure 1*) for applying a predefined torque of 0Nm, 1Nm and 4Nm on the ankle joint *(for more details, supplementary III)*. In the baseline condition (Baseline, 0Nm), the foot was placed in the set-up and was hanging down freely at the end of a triangular pillow. This baseline condition was always performed first, and was followed by a second and third condition, whereby 1Nm and 4 Nm ankle dorsiflexion torque was applied on the ankle, respectively (*supplementary III, Figure 2*). In each condition, the muscle tendon, muscle belly and fascicle length was assessed. The 1Nm condition was chosen as a condition in between baseline and 4NM, in which the ankle joint was often around neutral, i.e. 0 degrees of dorsiflexion in most children. The condition 4Nm was similar to the ankle torque applied by Weide *et al*. (2015) and was in most cases the maximal tolerated range of motion towards dorsiflexion.

**Figure 2:**
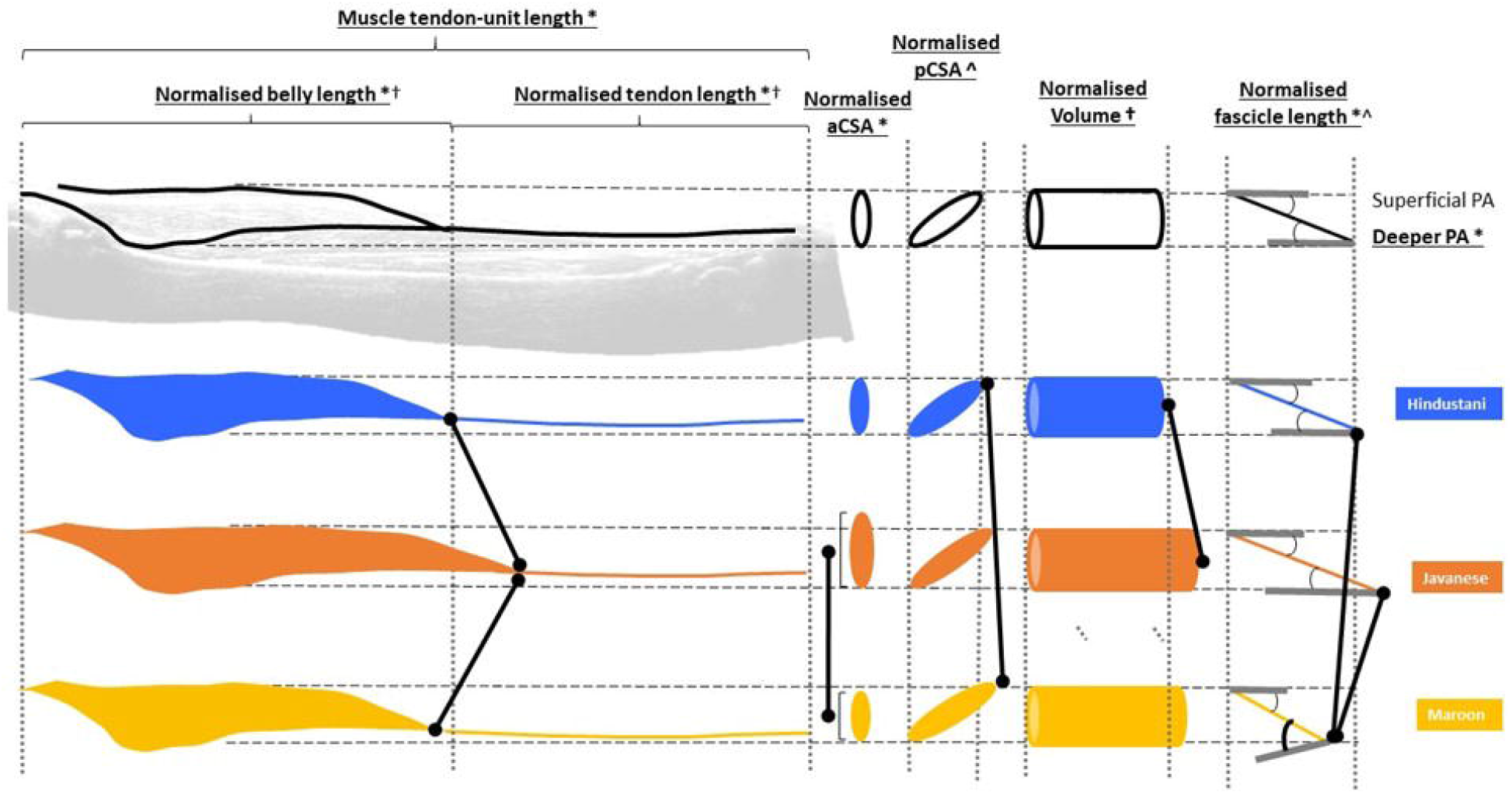
Assumed visualisation of the differences for each individual morphological and architectural parameter of the medial gastrocnemius for each ancestral group. Group differences *(p* < 0.05) are displayed with; * for significant difference between Maroon and Javanese children; ^ for significant difference between Maroon and Hindustani children; † for significant difference between Hindustani and Javanese children.

### Muscle strength

Isometric plantar flexion strength was measured with a MicroFed dynamometer (Hogan Health Industries, Inc. 8020 South 1300 West, West Jordan, USA) held manually by the assessor at 75% of the foot length. A mean of the maximal force (N) of three trials was used for further analysis and multiplied by 75% of the foot length (expressed in meter, m) to calculate the ankle torque (expressed Nm). Body weight was used for normalisation of ankle torque *(for more details, supplementary IV)*

### Statistical analysis

Statistical analysis was performed in Jeffreys’s Amazing Statistics Program (JASP) version 0.14.1. The Pearson’s Chi-Squared test was performed to evaluate equal distribution of boys and girls between ancestral groups. Normal distribution of data was assessed with the Shapiro-Wilk test and visual inspection of the plots for each ancestral group and each individual parameter. The data were described by means and standard deviation (SD) in case they were normally distributed, or by medians and interquartile ranges (IQR) in case they were not normally distributed. When data was normally distributed, the one-way Anova was used to investigate differences between ancestral groups, while the Kruskall Wallis H test was used when the data of one of the ancestral groups was not normally distributed. If the one-way ANOVA or Kruskall Wallis H test was significant, a post hoc t-test or Mann-Whitney U test was used with a Bonferroni correction for multiple testing, to define differences between the three ancestral groups. A two-way repeated measures Anova was used to define if there were differences in the extensibility of the muscle tendon, belly and fascicle between ancestral populations from baseline (0Nm) to 1NM and 4Nm. Level of significance, after correcting for multiple testing was set to *p* < 0.05.

## RESULTS

One hundred Hindustani (n=34), Javanese (n=34) and Maroon (n=32) children were included. Participants were distributed over the age-range 5 to 10 and Pearson’s Chi-Square (X^2^ (2, N = 100) = 0.602, *p* = .740) showed equal distributions of boys and girls between ancestral groups (supplementary table I).

There was a difference between ancestral groups for height (*p* = 0.023) and lower leg length (*p* = 0.003) (table 1). The post hoc pairwise group comparisons showed that the Maroon children were taller (*p* = 0.011) compared to Hindustani children, with a longer lower leg length compared to Hindustani (*p* = 0.007) and Javanese children (*p* = 0.003). Lower leg circumference, absolute and normalized plantarflexion strength were not different between groups (all *p* values ≥ 0.055).

**Table 1:** Group data for the anthropometric parameters of each ethnic group

|  | Hindustani (n=34) | Javanese (n=34) | Maroon (n=32) |  |
| --- | --- | --- | --- | --- |
|  | Median (IQR) | Median (IQR) | Median (IQR) | p-value |
| Age (years) | 7.58 (2.8) | 7.38 (2.6) | 8.64(2.8) | 0.168 |
| Height (cm) | 126.75 (15.6) | 121.00 (19.2) | 133.50 (17.4) | **0.023\*** |
| Body weight (kg) | 21.75 (7.5) | 22.25 (12.3) | 26.75 (11.8) | 0.224 |
| Lower leg length (cm) | 30.75 (5.3) | 29.50 (4.8) | 34.00 (5.1) | **0.003\*^** |
| Lower leg circumference (cm) | 23.50 (4.0) | 24.25 (6.5) | 25.00 (4.1) | 0.292 |
| Plantarflexion torque (Nm) | 11.13 (5.2) | 11.03 (5.6) | 14.84 (6.8) | 0.055 |
| Normalised plantarflexion torque (Nm/kg) | 0.44 (0.2) | 0.41 (0.2) | 0.51 (0.1) | 0.122 |

Abbreviations: IQR, interquartile range. Group differences ( $p < 0.05$ ) are displayed with; \* for significant difference between Maroon en Javanese children; ^ for significant difference between Maroon en Hindustani children; † for significant difference between Hindustani en Javanese children.

The MTU length was different between groups (*p* = 0.001), with significantly longer MTU length in Maroon compared to Javanese children (*p* = <0.001) (table 2). The belly length and tendon length, normalized to MTU-length, were different between groups (*p* = <0.001). The belly length was 11% shorter in Maroon and 7% shorter in Hindustani children compared to Javanese children (*p* = <0.001 and *p* = 0.002, respectively). Consequently, the tendon length was 11% longer in Maroon and 7% longer in Hindustani children compared to Javanese children (*p* = <0.001 and *p* = 0.002, respectively), while there were differences in muscle belly and tendon length between Hindustani and Maroon children (Figure 1). The normalized fascicle length of Javanese children was 23% longer compared to Maroon and 11% longer compared to Hindustani children (*p* < 0.001 and *p* = 0.010, respectively). The normalized muscle volume was different between groups (*p* = 0.007) and significantly higher in Javanese children compared to Hindustani children (*p* = 0.002). The normalized aCSA was higher in Javanese children compared to the other ancestral groups, with a significant difference compared to the Maroon children (*p* = 0.008). In contrast, the normalized pCSA was higher for Maroon children compared to the other ancestral groups, with a significant difference compared to the Hindustani children (*p* = 0.007). The pennation angle of the fascicle with the deeper aponeuroses showed differences between ancestral groups (*p* = 0.019) with a greater pennation angle in Maroon children compared to Javanese children (*p* = 0.015). Figure 2 shows a visualisation of the MG per ancestral group.

**Table 2:** Group data for the morphological parameters per ethnic group in resting position

|  | Hindustani (n=34)<br>Median (IQR) /<br>Mean (SD) | Javanese (n=34)<br>Median (IQR) /<br>Mean (SD) | Maroon (n=32)<br>Median (IQR) /<br>Mean (SD) | p-value |
| --- | --- | --- | --- | --- |
| Δ MTU length (cm) | 30.74 (4.6) | 28.90 (5.2) | 33.09 (5.6) | **<0.001** *^ |
| \| Muscle belly length (cm) | 16.23 (1.7) | 16.71 (21.4) | 16.97 (1.7) | 0.275 |
| \| Normalized muscle belly length | 0.53 (0.04) | 0.57 (0.04) | 0.51 (0.05) | **<0.001** *† |
| \| Tendon length (cm) | 14.59 (2.1) | 12.98 (2.4) | 16.23 (3.0) | **<0.001** *^† |
| \| Normalized tendon length | 0.47 (0.04) | 0.44 (0.04) | 0.49 (0.05) | **<0.001** *† |
| Δ Fascicle length (cm) | 4.00 (0.8) | 4.27 (0.6) | 3.67 (0.7) | **<0.001** *† |
| \| Normalized fascicle length | 0.24 (0.04) | 0.26 (0.03) | 0.22 (0.04) | **<0.001** *^ |
| Δ aCSA (cm<sup>2</sup>) | 4.68 (2.5) | 5.31 (1.5) | 5.09 (1.7) | 0.815 |
| Δ Normalized aCSA | 0.20 (0.04) | 0.22 (0.06) | 0.19 (0.05) | **0.020** * |
| Δ pCSA (cm<sup>2</sup>) | 12.05 (5.8) | 12.84 (6.1) | 15.85 (6.4) | **0.009** *^ |
| Δ Normalized pCSA | 0.53 (0.11) | 0.54 (0.10) | 0.59 (0.12) | **0.013**^ |
| Δ Volume (ml) | 46.98 (29.1) | 56.32 (32.1) | 61.22 (29.9) | 0.087 |
| Δ Normalized volume (ml/kg) | 2.16 (0.4) | 2.43 (0.4) | 2.32 (0.5) | **0.007**† |
| \| Superficial pennation angle (degrees) | 18.92 (3.6) | 18.52 (2.9) | 19.55 (3.7) | 0.466 |
| \| Deeper pennation angle (degrees) | 19.60 (3.3) | 18.71 (2.7) | 20.93 (3.4) | **0.019** * |

Abbreviations: SD, standard deviation; IQR, interquartile range; MTU, muscle-tendon unit; aCSA, anatomical cross-sectional area; pCSA, physiological cross-sectional area. Group differences ( $p < 0.05$ ) are displayed with; \* for significant difference between Maroon en Javanese children; ^ for significant difference between Maroon en Hindustani children; † for significant difference between Hindustani en Javanese children, | data was parametric analysis: (mean (SD)) are given en and p-values was were calculated by one-way Anova and post hoc independent t-tests), Δ non-parametric analysis: median (IQR) are given and p-values were calculated by Kruskal Wallis H and post hoc Mann-Whitney U test.

### Extensibility

The most frequently recorded ankle angle for the position at 0Nm was 15 degrees of plantarflexion in Hindustani children and 10 degrees of plantarflexion in Maroon and Javanese children. In the position of 1Nm, the most common measured ankle angle was 0 degrees in Hindustani children and 5 degrees dorsiflexion in the Maroon and Javanese children. The increase of ankle dorsiflexion from 1Nm to 4Nm, based on the most common recorded ankle angle, was 35 degrees in Hindustani children, 25 degrees in Javanese children and 20 degrees in Maroon children. Muscle belly, tendon and fascicle length significantly increased and pennation angle significantly decreased when applying increasing torques from baseline (0Nm) to 1Nm and 4Nm (all *p* values <0.001). Muscle belly length showed an overall difference between groups (*p* = 0.030), but the post hoc pairwise group comparisons showed no significant differences between specific ancestral groups. There were also no differences found in the extensibility of the muscle tendon (*p* = 0.482) and fascicle (*p* = 0.506) between ancestral groups (Figure 3).

**Figure 3:**
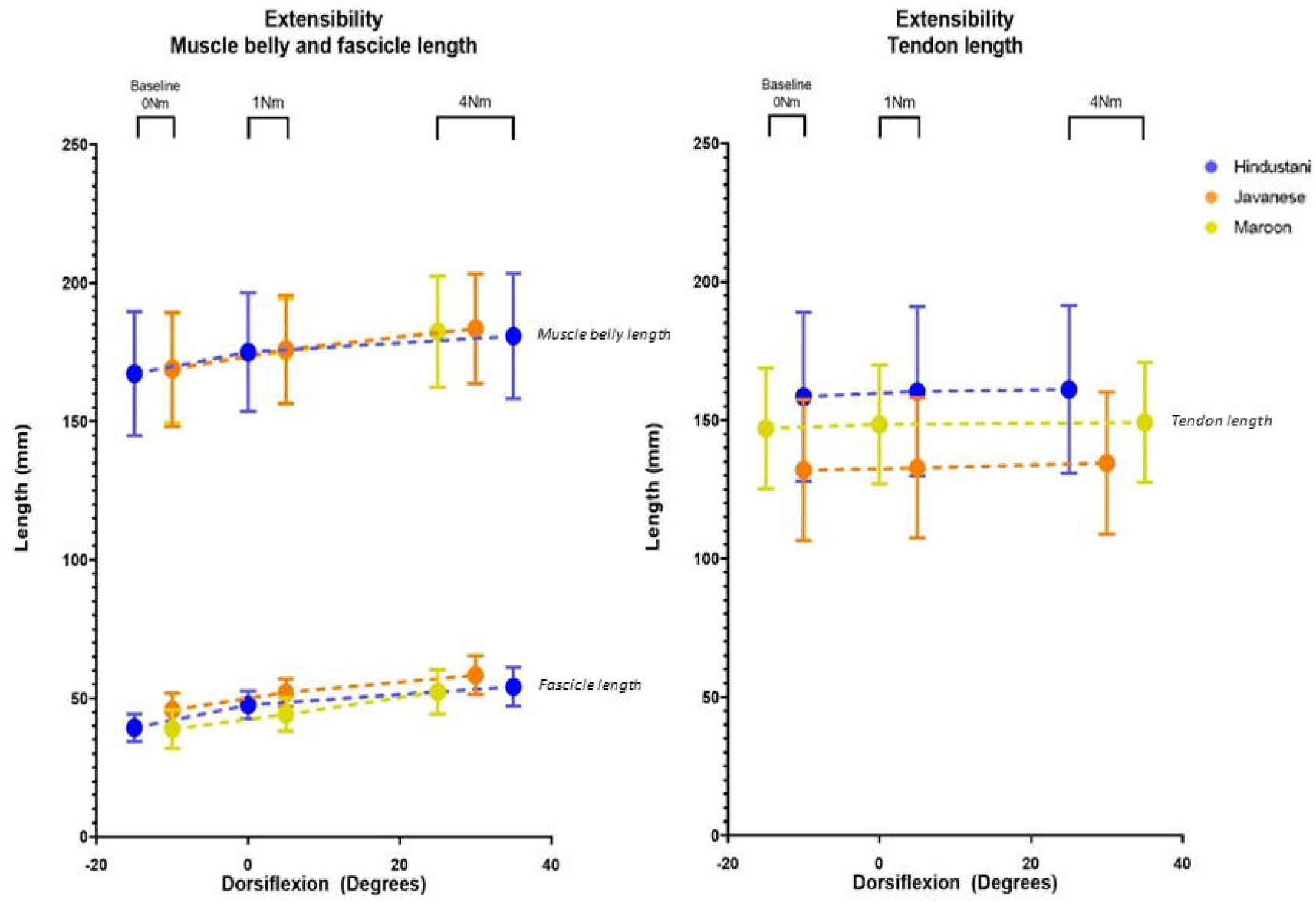
Extensibility of the muscle belly and fascicles (left graph) and tendon (right graph) from baseline (0Nm) to 1Nm and 4Nm. The mean and standard deviation are shown for each condition and for each ancestral group. The mean and standard deviation are plotted on the x-axis on the most frequent measured range of dorsiflexion for that specific condition and ancestral group.

## DISCUSSION

This study showed that there are differences in MG morphology (muscle belly-, tendon length and muscle volume) and architecture (fascicle length and deeper pennation angle) between children from three investigated ancestral groups, namely Maroon (Ghana, African), Hindustani (India, South Asian) and Javanese (Indonesia, Southeast Asian). These differences were present between all groups, but were most pronounced in Javanese children compared to Maroon or Hindustani children. There were no differences in the extensibility of the muscle belly, tendon and fascicles between ancestral groups. These findings highlight the need to take ancestry into account when comparing muscle morphology and architecture of the MG in children from different countries and to use ancestry-specific norm-databases when studying alterations in these intrinsic muscle properties in children with neurological or neuromuscular pathologies. This is the first study that investigated all macroscopic morphological and architectural parameters for the MG, as well as the extensibility of the muscle and tendon of the MG in one extended cohort of typically developing children, stratified in ancestral subgroups.

Differences were found in height and lower leg length, while no differences were found in body weight between the ancestral groups. Maroon (African) children were significantly taller with longer lower leg lengths compared to Javanese (Southeast Asian) and Hindustani (South Asian) children. Similar results were found in a recent study where lower leg length in Kenyan (African) children was longer compared to Japanese (East Asian) children, while there was no difference in body weight (Kunimasa *et al*., 2022). Lee *et al*. (2014) reported that African and Caribbean children (living in London) aged 5-10 years old tended to be taller, heavier, with larger leg circumferences and relatively longer legs compared to other ancestral groups, including South Asian children (Lee *et al*., 2014). While Lee *et al*. (2014) investigated similar age groups, current data showed no differences in body weight and calf circumference between ancestral groups. Longer leg lengths were also found in a recent study of Kunimasa *et al*. (2022) in children with African compared to East Asian ancestry. Therefore, it was considered important to normalize muscle length parameters when comparing muscle morphology and architecture between ancestral groups, especially when including children with African ancestry.

Normalized tendon length of the MG muscle was 7-11% longer in both Hindustani (South Asian) and Marron (African) children, compared to Javanese Southeast Asian) children. This is in line with a recent study, where differences in tendon length between Japanese (East Asian) and Kenyan (African) children were found, already at the age of 4 years old (Kunimasa *et al*., 2022). Similar percentages of difference in Achilles tendon length (i.e., ≈10% longer tendon lengths) between Kenyan (African) compared to Japanese (East Asian) adult runners has been reported (Kunimasa *et al*., 2014; Sano *et al*., 2015). The current study results revealed that the difference in tendon length between south Asian and African descent compared to southeast Asian descent exceeds the lower limit of difference (i.e., ≈4%) found in tendon length between TD and SCP (Barrett and Lichtwark, 2010; Kruse *et al*., 2021). Therefore, when comparing the tendon length of a group of TD and SCP children including different ancestral groups (especially including the southeast Asian ancestry), group-matching needs to take ancestral-variance into account.

Normalized muscle volume and aCSA was significantly larger between Javanese children compared to Hindustani children (for normalized muscle volume) and Maroon children (for normalized aCSA). A greater muscle volume could have been expected in Javanese children, because of their longer muscle belly length. On the other hand, no difference in normalized volume was found between Maroon and Javanese children, while the muscle belly length was significantly shorter in Maroon compared to Javanese children. Another contraction is the higher normalized pCSA in Maroon children compared to the other ancestral groups. Yet, so far, no other studies have evaluated the MG volume between children with different ancestry. While no differences in lower leg circumference between ancestries were found in current study, Refsdal *et al*. (2017) described differences at different shank heights between adults with African and Caucasian ancestry, which could suggest differences in the shape of the shank. Also the contradictory combination of differences in normalized muscle volume and muscle belly length found in the current study, suggest that children with different ancestry may have a different shape of the MG muscle belly. This could be further explored using statistical shape modeling in future studies (Ghouth *et al*., 2022).

The muscle belly length of the MG is defined by the muscle architectural parameters fascicle length and pennation angle. Normalized fascicle length was lower for Maroon compared to Hindustani and Javanese children, while the deeper pennation angle was greater in Maroon children compared to Javanese children. A deeper pennation angle was also found in a recent study of Kunimasa *et al*. (2022) for children with African compared to East Asian ancestry. Despite the fact that the deeper pennation angle is considered more sensible for errors than the superficial pennation angle due to the unavoidable tilt of the probe during the acquisition (Bénard *et al*., 2009), the differences in the deeper pennation angle were between ancestral groups. This may strengthen our earlier assumption of muscle shape differences between children with different ancestral background. The results for fascicle length were in line to the findings of Kunimasa *et al*. (2022), who also reported longer normalized fascicle lengths in Kenyan children compared to Japanese children. In short, fascicle length was the longest in Javanese children and smallest in Maroon children, which is in line with a longer belly length in Javanese compared to Maroon children.

The significant increase in muscle belly, tendon and fascicle length and significant decrease in pennation angle when a torque of 1 and 4Nm was applied compared to the condition of 0Nm torque, are in line with the changes found during passive stretching of the MG (Bolsterlee *et al*., 2017; Kalkman *et al*., 2018). According to models that present the stiffness caused by the arrangement of muscle fascicles (and thereby the sarcomeres) (Kruse *et al*., 2021), the increase in fascicle length is assumed to decrease the passive stiffness and thereby increase the extensibility. Although fascicle length was significant different between ancestral groups, we found no differences in the extensibility of the muscle belly, muscle tendon and fascicles between ancestral groups. This suggests that other components, such as intramuscular connective tissue (Järvinen *et al*., 2002), may have contributed to the observed extensibility.

We did not expect to find great differences in extensibility of the tendon between typically developing children with different ancestry. Weide *et al*. (2020) found no differences in extensibility of the MTU from baseline to 4Nm between TD and SCP children, which was in contrast to other studies that found less muscle belly and more muscle tendon extensibility in SCP compared to TD children (Theis, Mohagheghi and Korff, 2016; Kalkman *et al*., 2018; Weide *et al*., 2020). The contradicting results could have been caused by difference in methods. The current study and Weide *et al*. (2020) measured the extensibility statically and the other studies defined extensibility during dynamic stretching. However, the current study data created a unique norm-database for the extensibility of the muscle belly, muscle tendon and fascicle length that can be used as a reference when investigating the extensibility parameters of SCP children.

Studies in adults have indicated differences in the muscle morphology between Caucasian, African and East Asian descendants. One study reported smaller muscle thickness, shorter fascicle length and longer tendon length of the MG in African compared to Caucasian descendants (Refsdal, 2017). When comparing African descendants to East Asian descendants, shorter fascicle lengths, greater pennation angles and longer Achilles tendon lengths were found in African descendants compared to East Asian descendants (Sano *et al*., 2015; Kunimasa *et al*., 2022). These results are in line with the current findings. It remains unknown if the differences found between the ancestral groups in Suriname can be generalized to ancestral groups in other countries. We advise that future studies report the ancestral background when describing muscle morphology of children and adults and that ancestral specifications are well-considered when establishing and using normative databases.

## Limitations

Besides ancestral differences, also nutritional and other environmental factors can influence muscle growth. Therefore, we included only children living in the capital city of Paramaribo to minimalize cultural differences between ancestral groups (e.g. beliefs and traditional food) and living circumstances (e.g. housing, possibility of schools, access to sports clubs). Although we do not expect great differences by these factors from 5-11 years old, it would be interesting to take these factors in account in further studies or studies with older children.

This study did not focus on the relation of muscle morphology to function and performance, while the moments arm at the joints can differ between ancestral groups (Hanson *et al*., 1999; Kunimasa *et al*., 2022). This is of interest for further research, since ancestry-specific features of the MG muscle morphology may potentially be related to force production and performance in walking and running (McCarthy *et al*., 2006; Sano *et al*., 2014; Butler and Dominy, 2016).

## Conclusion

The current study showed differences in MG morphology (muscle belly-, tendon length and muscle volume, aCSA) and architecture (pCSA, fascicle length and deeper pennation angle) between children with different ancestry, the Maroon (Ghana, African), Hindustani (India, South Asian) and Javanese (Indonesia, Southeast Asian), aged 5 to 10 years old. These differences were present between all groups, but were most pronounced in Javanese children compared to Maroon or Hindustani children.

The current results revealed that ancestry-specific reference data are needed when investigating altered muscle morphology in neurological or neuromuscular pathologies, such as cerebral palsy. We recommend that future studies report the ancestral background when describing muscle morphology and architecture of children and that ancestral specifications in normative databases should be included.

## Supporting information

supplementary III

## Data Availability

All data produced in the present study are available upon reasonable request to the authors

## Notes

### Competing Interest Statement

The authors have declared no competing interest.

### Funding Statement

The authors are supported by the following funding bodies: the Flemish Research Foundation (FWO), Belgium, TBM project TAMTA, grant number T005416N, the Flemish Research Foundation (FWO), Belgium, research grant number G0B4619N and the internal KU Leuven research grant 3D-MMAP, Belgium, C24/18/103. The authors wish to thank all participants in this investigation and all students involved in recruitment and assistance during measurements.

### Author Declarations

The Ethics Committee (Commissie van Wetenschappelijk Onderzoek) of the Ministry of Health in Suriname gave ethical approval for this work (VG 003-17)

