## supplementary I for "Muscle morphology and architecture of the medial gastrocnemius between typically developing children with different ancestral backgrounds"

**Anthropometric measurements**

Anthropometric measurements included body height and weight, lower leg length and calf circumference. Body height was measured with a SECA 213 stadiometer (recorded in centimeters) and body weight was defined using a calibrated SECA 762 mechanical personal scale (Medical Measuring Systems and Scales, Chino, CA, USA) rounded up to the nearest 0.5 kilograms. Lower leg length was measured in supine lying position with the knee in full extension and the leg in line with the trunk by use of a measuring tape (recorded in millimeters) and was defined as the distance between the lateral epicondyle of the femur and the distal tip of the lateral malleolus of the fibula (Baumbach *et al.*, 2014). Calf circumference was measured in prone lying position at 50 percent of the muscle belly length. The 50 percent was defined as half of the distance from the epicondyle of the femur to the muscle tendon junction, which was identified by ultrasound and marked on the leg. Calf circumference was measured with the leg in relaxed position without contraction of the plantar flexors, perpendicular to the long axis of the calf and tight enough to be in contact with the skin without compression rounded up to the nearest 0.5cm.

**Range of motion**

Maximal passive range of motion for ankle dorsiflexion was measured for the children using a universal goniometer (7" Baseline Plastic goniometer, Fabrication Enterprises). The maximal passive dorsiflexion was assessed in a supine lying position with the knee maximally extended. To remove the full tension of the soleus muscle, a second test was performed with the knee and hip in 90^o^ of flexion (Mudge *et al.*, 2014). In all measurements, the fulcrum of the goniometer was placed on the lateral malleolus, the stationary shaft was held parallel to the long axis of the fibula and the movable shaft was kept parallel to the long axis of the fifth metatarsal bone (Mudge *et al.*, 2014). During both measurements, the hind foot was held in neutral position (no varus or valgus was allowed).
