## supplementary II for "Muscle morphology and architecture of the medial gastrocnemius between typically developing children with different ancestral backgrounds"

**2D freehand ultrasound**

A conventional B-mode 2D ultrasound (Telemed EchoBlaster128, Vilnius, Lithuania) was used for all ultrasound measurements. The system included an echo blaster with 128 beamformer (cext-1z REV:C, serial: 2311-160818-0320) and a linear transducer with a 55 mm field of view (HL9.0/60/128Z-2, scanning frequency: 5.0-12.0 Mhz, serial: 31027-160823-0011). Data collection was performed using STRADWIN software with a frequency of 30hz. For standardization of the 2D ultrasound measurements, two lines were drawn on the leg of the participant (supplementary II, Figure 1a). One line was drawn transverse to the leg at 50 percent of the muscle belly length measured from the medial epicondyle of the femur to the muscle tendon junction of the medial muscle belly. Another line was drawn parallel to leg length at 50 percent from the medial muscle belly border to the lateral muscle belly border. The crossing point of the transverse and longitudinal lines were used for standardized placement of the transducer for the 2D US measurements that were acquired to define fascicle length, pennation angle and anatomical cross sectional area (aCSA).

*Fascicle length and pennation angle*

Fascicle length and pennation angle were defined based on 2D ultrasound imaging as described above. The long side of the transducer was positioned on the longitudinal line that was drawn at 50 percent of the belly length. The mid-point of the transducer was positioned at the crossing point of the two marked lines and ultrasound images are made while the transducer was tilted back and forth with the transducer in contact with de drawn longitudinal line (close to the axis of rotation) (supplementary II, Figure 1b). To ensure that the transducer stayed in contact with the leg while the transducer was tilted, one hand of the investigator held the transducer at the long side at the axis of rotation and the other hand of the investigator tilted the transducer medial and lateral. Data analysis was performed in one picture from the movie where both aponeuroses were clearly visible. The fascicle length was measured from the deeper to the superficial aponeuroses (in cm). The pennation angle was recorded as the angle of the fascicle with the deeper aponeuroses (in degrees), the deeper pennation angle, and as the angle of the fascicle with the superficial aponeurosis, the superficial pennation angle (also in degrees). Fascicle length, superficial and deeper pennation angles were extracted from the same image with a custom-made matlab software (Mathworks, R2017b).

| 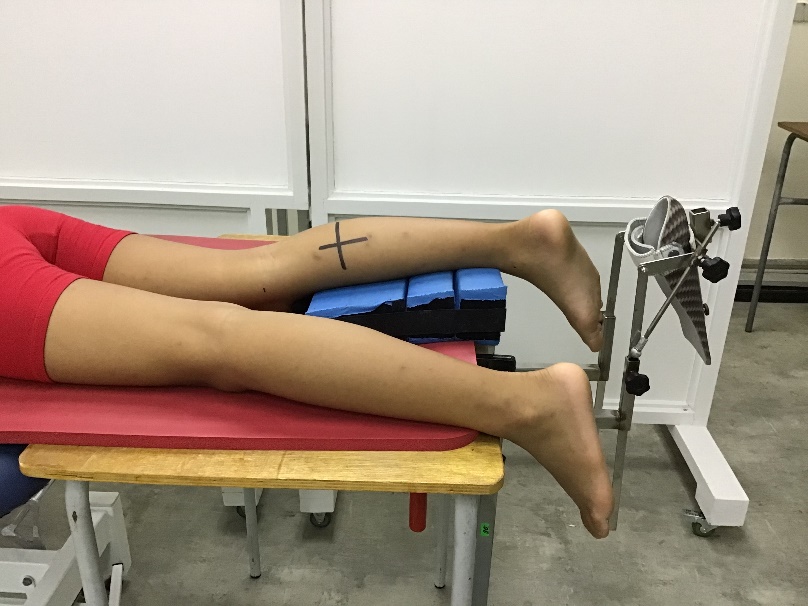 | 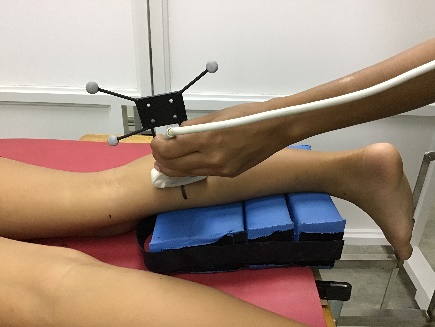 | 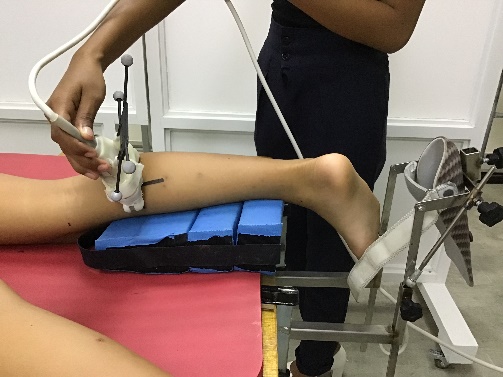 |
| --- | --- | --- |

Supplementary II, Figure 1a (left), 1b (middle), 1c (right): position of the ultrasound transducer for 2D freehand ultrasound

*Anatomical cross-sectional area*

The aCSA was measured at 50 percent of the muscle belly length on the transverse line (supplementary II, Figure 1c). The mid-point of the transducer was positioned at the crossing point of the drawn transverse and longitudinal line. During the 2D ultrasound images, the transducer was tilted a few degrees up and down while the transducer was kept in contact with de drawn transverse line (the axis of rotation). To ensure that the transducer stayed in contact with the leg while the transducer was tilted, one hand of the investigator stabilized the transducer at the long side at the axis of rotation and the other hand of the investigator tilted the transducer back and forth. Data analysis was performed in the 2D ultrasound image wherein both the upper and lower aponeurosis were clearly visible, to ensure that the transducer was perpendicular positioned to the muscle. If the aCSA was wider than 5cm (the width of the transducer) and therefore not visible in one ultrasound image, the CSA was distracted from the combination of two images of a multiple sweep. The aCSA was calculated by manually outlining the borders of the medial gastrocnemius using of STRADWIN software (version 6.0; Mechanical Engineering, Cambridge University, Cambridge, UK) and normalised by body weight.

**3D freehand ultrasound**

For the 3D freehand ultrasound measurement, the ultrasound system was combined with an OptiTrack V120: Trio camera system with 3 cameras and a 1 mm spatial resolution (serial: #330030) to track four passive markers that were attached to the transducer (supplementary II, Figure 2). This method was shown to be reliable and valid to measure muscle volume, belly length, tendon length and muscle-tendon unit (MTU) length (Cenni *et al.*, 2016). An Aquaflex ultrasound concave-shaped portico with gel pad was used for optimal contact and signal transmission between the skin and the transducer of the ultrasound. The depth of the curve of the gel pad was determined depending on the size of the participant’s lower leg*.* Cenni et al. 2018 demonstrated that the use of a portico and gel pad is a more reliable and valid method to measure muscle volume compared to the approach with direct contact of the transducer on the skin (Cenni *et al.*, 2018). Ultrasound Aquaflex gel was used to attach the gel pad to the transducer and additionally the same gel was used as a conductive medium between the skin and gel pad.

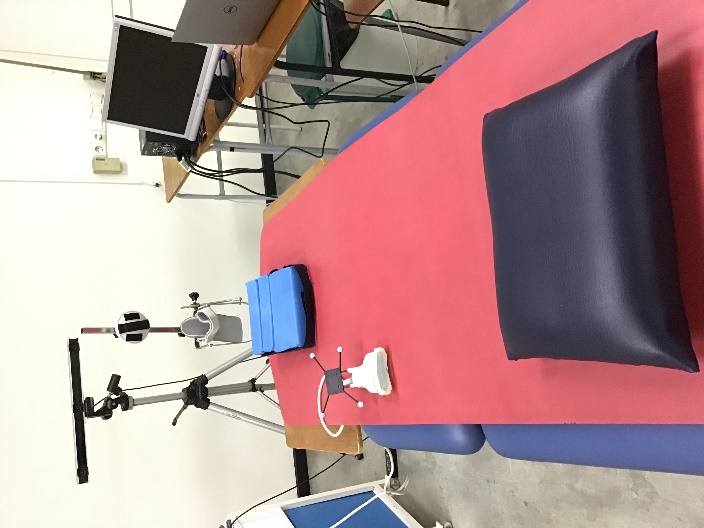

Supplementary II, Figure 2: The 3D freehand ultrasound measurement set-up.

*Muscle, tendon and MTU length*

Ultrasound images of the muscle and tendon of the medial gastrocnemius were acquired in a transverse orientation in two separate sweeps. Each sweep was made from just above the femur condyles over the muscle belly and tendon until the distal end of the calcaneus. Belly length was defined as the distance between the most superficial point of the medial femur condyle and the first image where no muscle tissue of the medial gastrocnemius was visible, further labelled as the muscle tendon junction (MTJ). Tendon length was defined from MTJ to the most proximal point on the calcaneus representing the tendon insertion. MTU length was recorded as the sum of belly and tendon length. If there were missing data or movements of the child for the first sweep, the second sweep was used for defining the lengths. Both muscle belly and tendon length were normalized to lower leg length and MTU-length.

*Muscle volume*

For muscle volume, acquisitions of the medial gastrocnemius were taken while moving the transducer in a transverse position, from proximal of the femoral condyles to the distal end of the calcaneus. The ultrasound transducer was moved perpendicularly to the surface of the lower leg at an almost constant velocity, while 30 ultrasound images per second were recorded. If the muscle was too wide to fit in an ultrasound image, a multiple sweep, consisting of two separate sweeps that were afterwards combined, was acquired. Muscle volume was calculated by manually outlining the borders of the medial gastrocnemius starting at the most proximal edge of the medial condyle of the tibia, and ending at the muscle tendon junction. Muscle segmentations were made each centimeter and a linear interpolation between the outlined borders was applied to compute the muscle volume (in ml). In addition, muscle volume was normalized to body mass (ml/kg).

*Physiological cross-sectional area*

Physiological cross-sectional area (pCSA) was calculated similar to other studies by the following equation; pCSA = (muscle volume * cosinus (superficial pennation angle))/fascicle length (Barber *et al.*, 2011; Herskind *et al.*, 2016). The muscle volume, superficial pennation angle and fascicle length were obtained as described in the paragraphs above.
