## supplementary III for "Muscle morphology and architecture of the medial gastrocnemius between typically developing children with different ancestral backgrounds"

**Measurement set up**

The muscle morphology was measured in prone position with the hip in extension. The lower leg was placed on an triangular pillow that was adjustable to support different lower leg lengths, resulting in 30 degrees of knee flexion during all measurement conditions. This 30 degrees of knee flexion was chosen to allow that this method could also be used in children with SCP who have limited knee extension. In line with most studies in literature, the muscle morphology was first measured in a neutral or resting position of the ankle (Hanssen *et al.*, 2021; Williams *et al.*, 2021). Besides this neutral or resting position, the muscle morphology was also measured in a stretched condition by a applying a predefined torque on the ankle joint. A custom made set-up, with the foot of the child placed in the orthosis on which a force sensor was fixed, allowed the investigator to apply different torques on the ankle joint. Three different sizes (small, medium, large) of orthoses were available to ensure that the rotation axis of the set-up could be visually aligned with the axis through the malleoli (red dot; supplementary III, Figure 1). A dynamometer was attached by an iron bar to the footplate of the orthosis. The amount of force (green arrow in picture 1) that was applied by the investigator on the dynamometer (MicroFed2, Hogan Health Industries, Inc. 8020 South 1300 West, West Jordan, USA) was calculated by dividing the desired torque by the moment-arm (yellow line, supplementary III, Figure 1) of the dynamometer with respect to the ankle joint (red dot, supplementary III, Figure 1). When the desired ankle torque was gained (i.e., 0Nm, 1Nm, or 4Nm), the axis of the custom made set-up was fixed to that certain position, and the muscle morphology measurements were performed in this static position (*supplementary III, Figure 2)*.

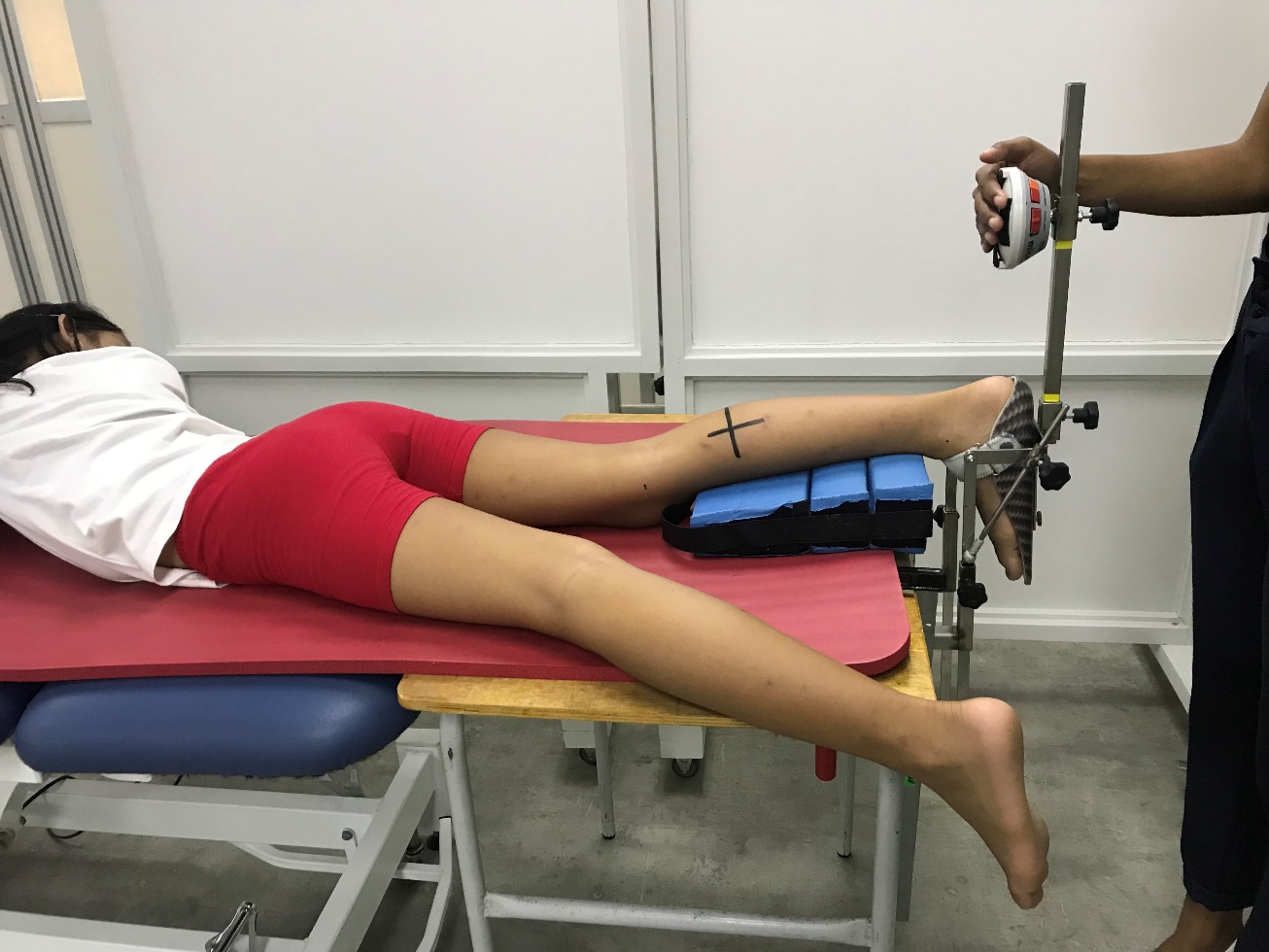

Supplementary III, Figure 1: The set-up for fixation of the ankle joint by applying different torques

RP	Baseline	1Nm	4Nm

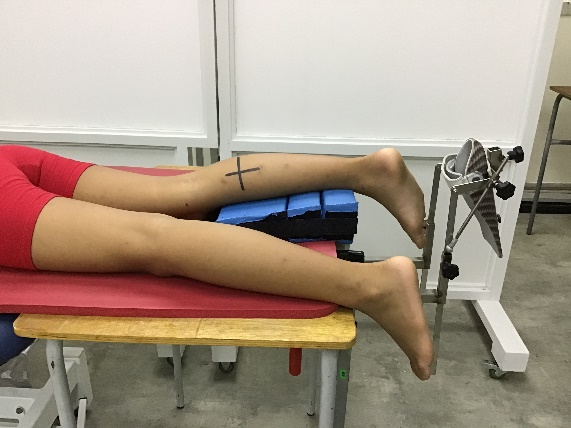

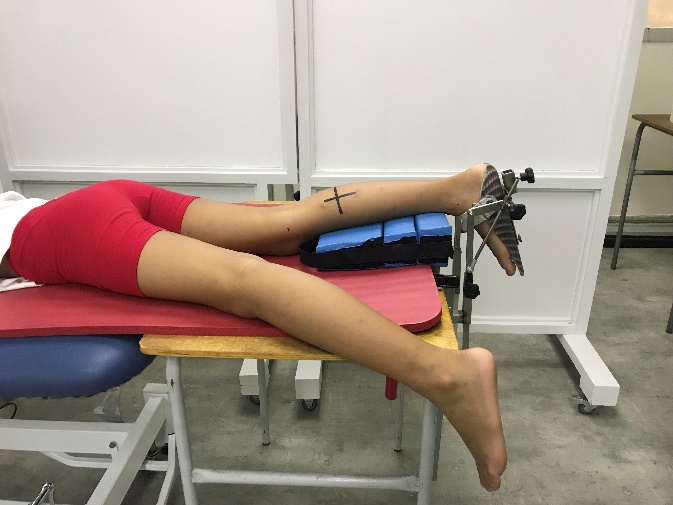

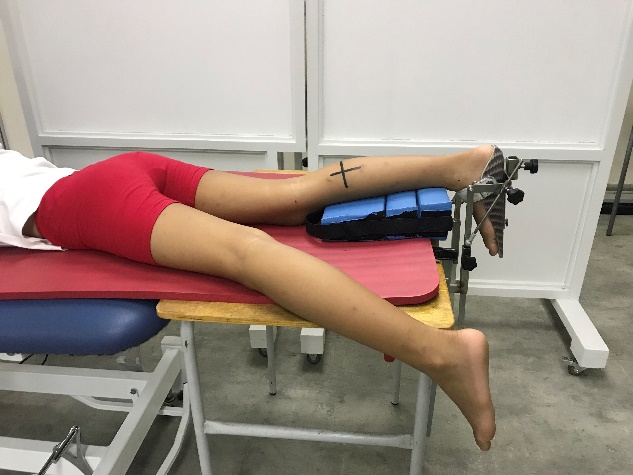

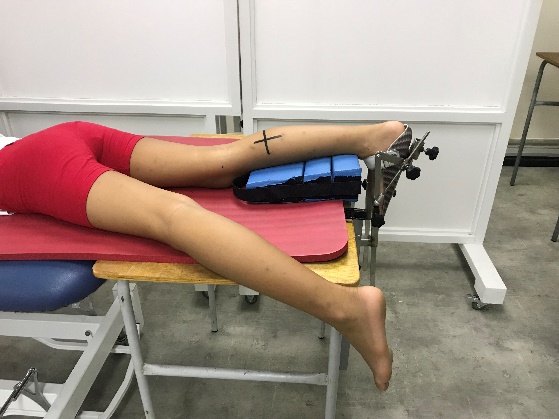

Supplementary III, Figure 2: Position of the ankle during the different conditions; resting position, baseline (0Nm), 1Nm and 4Nm.
