## supplementary IV for "Muscle morphology and architecture of the medial gastrocnemius between typically developing children with different ancestral backgrounds"

**Muscle strength**

Isometric plantar flexion strength was measured with the foot placed in a small light-weight orthosis with a rigid footplate with the child lying in prone and the ankle joint in the rest position similar to the ultrasonography measurements. A MicroFed dynamometer (Hogan Health Industries, Inc. 8020 South 1300 West, West Jordan, USA) was manually kept at 75 percent of the foot length. Foot length was measured from the lateral malleolus to metacarpal-phalangeal joint of the second toe. The isometric strength measurement was performed while the arms of the child arms were in relaxed position and the upper leg was fixed with a strap to the bed to avoid movement of the leg during measurement. The participant was first asked to relax their leg and foot while the investigator placed the dynamometer to the footplate at 75 percent of the total foot length. From the resting position of the ankle joint, the participant was asked to press the foot (fixed in the orthosis) to plantarflexion as much as possible, while the dynamometer was manually fixed by the investigator such that movement to plantarflexion was not possible (supplementary IV, Figure 1). The maximal isometric measurement was repeated 3 times and the maximum force for each of the three trials was recorded. The isometric muscle strength measurements were always performed after the ultrasound measurements. A mean of the maximal force (Newton) of the three trials was used for further analysis and multiplied by 75 percent of the foot length (expressed in meter) to calculate the ankle torque (expressed in Nm).

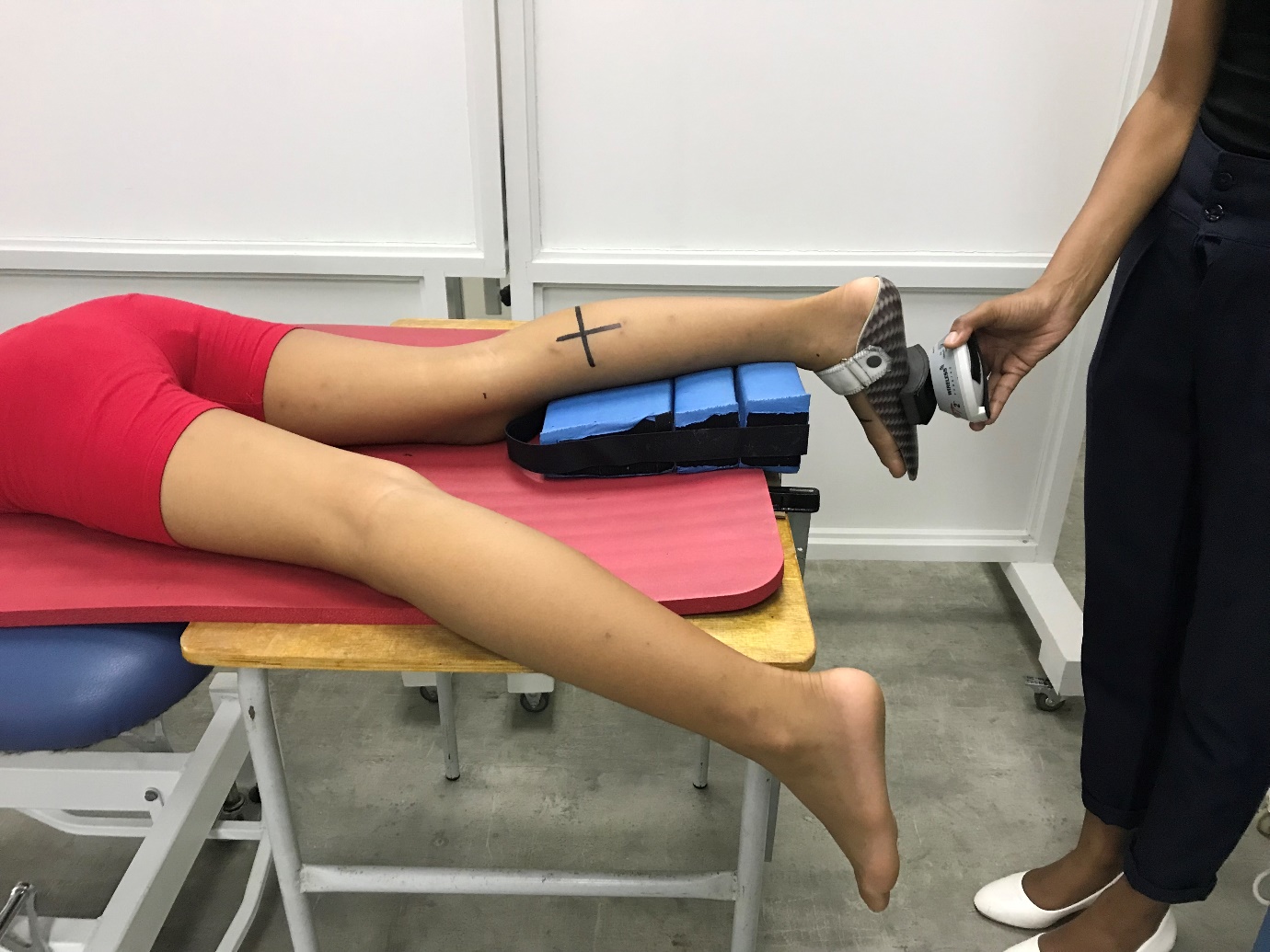

Supplementary IV, Figure 1: Manually fixation of the dynamometer by the investigator during plantar flexion strength measurement
