## supplementary table I for "Muscle morphology and architecture of the medial gastrocnemius between typically developing children with different ancestral backgrounds"

| **Age** | **H (n)** | **M (n)** | **J (n)** |
| --- | --- | --- | --- |
| 5 | 6 | 6 | 3 |
| 6 | 7 | 9 | 5 |
| 7 | 6 | 6 | 5 |
| 8 | 4 | 5 | 6 |
| 9 | 6 | 4 | 6 |
| 10 | 5 | 4 | 7 |
| **Gender (Boys/Girls)** | 15/19 | 17/17 | 13/19 |

De number (n) of Hindustani (H), Maroon (M) and Javanese (J) children that participated in the study per age category. Also the number of boys and girls that were included per ancestry are shown.
